# Early user experience and lessons learned using ultra-portable digital X-ray with computer-aided detection (DXR-CAD) products: A qualitative study from the perspective of healthcare providers

**DOI:** 10.1101/2022.11.04.22281963

**Authors:** Zhi Zhen Qin, Rachael Barrett, Maria del Mar Castro, Sarah Zaidi, Andrew J. Codlin, Jacob Creswell, Claudia M. Denkinger

## Abstract

**Background:** Recent technological and radiological advances have renewed interest in using X-rays to screen and triage people with tuberculosis (TB). The miniaturization of digital X-ray (DXR), combined with automatic interpretation using computer-aided detection (CAD) software can extend the reach of DXR screening interventions for TB. This qualitative study assessed early implementers’ experiences and lessons learned when using ultra-portable (UP) DXR systems integrated with CAD software to screen and triage TB.

**Methods:** Semi-structured interviews were conducted with project staff and healthcare workers at six pilot sites. Transcripts were coded and analyzed using a framework approach. The themes that emerged were subsequently organized and presented using the Consolidated Framework for Implementation Research (CFIR).

**Results:** There were 26 interviewees with varying roles: supervisory, clinicians, radiographers, and radiologists. Participants recognized the portability as the main advantage, but criticize that it involves several compromises on throughput, internet dependence, manoeuvrability and stability, as well as suitability for patients with larger body sizes. Furthermore, compared to use the hardware and software from the same supplier without digital health information system, complexity increases with interoperability between hardware and software and between different electronic health information systems. Currently, there is a limited capacity to implement these technologies, especially due to need for threshold selection, and lack of guidance on radiation protection suitable for UP DXR machines. Finally, the respondents stressed the importance of having protected means of sharing patient medical data, as well as comprehensive support and warranty plans.

**Conclusion:** Study findings suggest that UP DXR with CAD were overall well received to decentralize radiological assessment for TB, however, the improved portability involved programmatic compromises. Main barriers to the uptake included the insufficient capacity and lack of guidance on radiation protection suitable for UP DXR.

## Introduction

Tuberculosis (TB) is a transmissible bacteriological disease behind a global health crisis. Responsible for 1.6 million deaths in 2021 according to the World Health Organization (WHO)[1], TB kills more people every year than any other infectious disease despite the availability of effective treatments. The global TB burden is concentrated in lower middle-income countries, eight of which account for two thirds of incident TB [1]. COVID-19 has recently set back the international campaign to eliminate TB by at least five years [2]. However, even before the COVID-19 pandemic, the global campaign against TB was not on track to meet the targets set by the Global Plan to End TB [3].

The persistent gap between TB incidence and treatment notification has driven renewed interest in active screening for TB using chest X-ray (CXR) [4]. This change has taken place against a backdrop of a fundamental paradigm shift from TB control towards TB elimination and significant technological advances [5]. New detector panel technology has spurred innovation in digital X-ray (DXR) technology to produce smaller, battery-powered devices with reduced radiation emissions. These are designed to be portable in a suitcase a backpack, or even hand-held. Meanwhile, computer-aided detection (CAD) software can provide automatic and standardized interpretation of CXR in the absence of radiologists, who are often scarce in lower-income settings [6]. A recent landscape review identified 13 CAD products commercially available for TB detection, with new products and product versions in the pipeline [7]. Independent evaluations show CAD software can read CXR just as accurately as – or even better than – human readers [8, 9], and their use was recommended by WHO in the 2021 update to its TB screening guidelines [4]. Applying ultra-portable DXR with CAD software (UP DXR-CAD) offers an opportunity to address the barriers faced by CXR screening in the past, thereby improving TB detection and treatment.

To date, the CAD literature has focused primarily on the performance evaluation [8-11], and the experiences of early CAD implementers has not been well-described. This is important, as no matter how accurate the detection technology, if it is not acceptable to users and/or implementation barriers exist, the full potential of this novel technology will not be reached. Therefore, we conducted this qualitative study, through semi-structured interviews, to uncover the acceptability and perceived limitations of the new technology from the health provider perspective, including lessons learned and opportunities yet to be explored.

## Methods

### Ethics statement

This research was approved and monitored by Ethikkommission der Medizinischen Fakultät Heidelberg (reference number: S-399/2021) in accordance with consensus ethical principles derived from international guidelines, including the Declaration of Helsinki (7^th^ revision, 2013). The participants, all above 18 years of age, were informed that their participation in the study was voluntary, and that they could submit written or oral notification of withdrawal at any time, before or after the interview, without explanation or penalty. Written informed consent was obtained from all participants, with some wishing to be acknowledged as contributors and some remaining anonymous. The personal information of the participants and all other confidential information has been stored and secured according to the medical confidentiality and other provisions of the General Data Protection Regulation (GDPR), the German Federal Data Protection Act (BDSG) and the associated Baden-Württemberg State Data Protection Act (LDSG BW).

### Context and participants

At the time interviews, all implementation of DXR-CAD had been conducted in small scale pilot settings; none of the interviews correspond to a programmatic setting. Participants were identified among staff of projects known to be using DXR-CAD for TB screening. This was achieved through the network of in-country partners of the Stop TB Partnership’s TB REACH initiative in several high TB burden countries, including Cambodia, Nigeria, Pakistan, Uganda, Vietnam, and Zambia.

Six DXR-CAD pilot projects were identified. The projects in Nigeria and Vietnam deployed DXR-CAD to support community-based active TB case finding (ACF) in hard-to-reach areas, such as remote or mountainous areas and islands. The remaining four projects primarily employed a facility-based screening; in Zambia and Cambodia DXR-CAD was used in remote clinics, while in Pakistan these tools were used in tertiary hospitals, and in Ghana they were used for systematic screening in prisons. In both Cambodia and Pakistan, these tools were also used for community-based ACF in rural and mining areas, similar to the projects in Nigeria and Vietnam. Four ultra-portable DXR systems (Fujifilm FDR Xair, Delft Ultra, Delft Light, MINE2) and three CAD software products (Lunit INSIGHT CXR, qXR, CAD4TB) were used by the six projects in a range of combinations (Table 1). All the equipment was commercially available at the time of implementation.

**Table 1:** Project and participant Characteristics.

| Location | Pakistan | Vietnam | Zambia | Ghana | Nigeria | Cambodia |
| --- | --- | --- | --- | --- | --- | --- |
| Use case | Community ACF* and facility-based screening | Community ACF* | Facility-based screening | Facility-based screening | Community ACF* | Community ACF* and facility-based screening |
| Setting | Tertiary hospitals and mining communities | Hard-to-reach rural areas and islands | Remote clinics and door-to-door | Medium security prison | Semi-urban communities | Remote clinics and hard-to-reach areas |
| **Respondents** | Project Director (1), Radiologists (5) | Research Director (1), Senior Technical Advisor (1), Radiographer (1), Clinician (1) | Project Lead (1), Clinicians (2), Radiographer (1) | Clinician (1), Radiographer (1) | Project Coordinator (1), Executive Director (1), Senior Programme Manager (1), radiographer (1), screener (1) | Primary Investigator (1), Project Supervisor (1), Project Planner and Researcher (1), Clinicians (4). |
| **Length of piloting at time of interview** | 11 months | 4 months | 11 months | 2 days | 6 months | 5 months |
| **UP DXR device** | Fujifilm FDR Xair | Fujifilm FDR Xair | Fujifilm FDR Xair | Delft Ultra | Delft Light | MINE2 |
| **CAD software** | Lunit INSIGHT CXR | Lunit INSIGHT CXR and qXR | CAD4TB | CAD4TB | CAD4TB | Lunit INSIGHT CXR |
| **Internet requirement** | Offline (computer) | Offline (Lunit box)<br>Online (qXR computer) | Online (box) / Hybrid | Offline (box) | Online (box)/ hybrid | Offline (box) |

\*ACF: provider-initiated Active Case Finding

We interviewed representatives from the six projects following a purposive sampling approach to identify information-rich case studies that best reveal user experiences with DXR-CAD. To guide recruitment, we followed the concept of information power, whereby a targeted group of participants were recruited as they met the specific aims of gathering quality information from both health workers and programmatic personnel involved in the early adoption of DXR-CAD for TB screening and triage [12]. We sought to include at least two healthcare workers (HCWs) per site, including radiographers, radiography technicians, radiologists, and other clinicians involved in routine care provision. This was achieved for all sites, except Nigeria, where no HCWs were available. Interviews with HCWs included questions exploring their experience of use, regulations, and logistics (e.g., procurement, installation, etc.). To gain insight regarding programmatic considerations and implications, we also interviewed at least one program manager per site, given the limited number of eligible participants.

### Data collection

From April to June 2021, ZZQ and RB who were both associated with the Stop TB Partnership TB REACH team, conducted individual and small-group semi-structured interviews with a total of 17 participants. In addition to these interviews, written answers were collected by an additional 11 participants.

Each interview was conducted using Microsoft Teams, due to pandemic-related travel constraints. Lasting 60–90 minutes, the questions tackled the programmatic context, technical characteristics, user experience, manufacturer service, and training, relevant to both CAD and DXR. Extensive notes were taken during interviews by one interviewer and then transcribed verbatim from audio recordings. The interview topic list was revised several times to reflect the feedback and observations uncovered by the interview process. Interviews were conducted in English, except for the clinical staff from the pilot project in Vietnam. A questionnaire developed based on the topic list was sent to the participants through the program managers who translated the questions into the local language. The answers were collected verbally or by quick messaging tools in their local language and translated back to English by the respective program managers.

Questions that respondents could not answer in sufficient detail were forwarded to clinicians or radiologists from project field teams, who provided answers either by separate interview or in written form, depending on language requirements. Respondents were encouraged to provide supplementary photographs or documents to aid understanding. While respondents did not review transcripts, all respondents were followed-up by email to answer additional questions and for clarification, where necessary. Respondents were also given the opportunity to review and comment on the write-up of the results.

### Data analysis

All transcripts, interview notes, and follow up questionnaires, were coded, using both deductive codes from the topic list, and inductive codes emerging from the texts. Data analysis was conducted using Microsoft Excel by ZZQ, RB and MMC, and supervised by CMD and JC, following the five-stage framework approach outlined by Pope et al [13]. The two interviewers agreed on initial codes (Supplementary material, S. Table 1) following a review of the transcripts and then independently coded three transcripts using this initial coding framework. Following discussion, codes were refined and amalgamated to form the final framework. The coded material was organized into themes using thematic analysis [14-16]. Six themes were identified, with 22 secondary and tertiary codes. The team discussed the themes, and existing theories or frameworks that could guide presentation of the data. From this, the Consolidated Framework for Implementation Research (CFIR), a conceptual framework which aims to promote the systematic analysis of implementation studies [17], was used to organize and present findings as barriers and facilitators along the CFIR constructs of adaptability, complexity, inner setting (e.g., readiness for implementation, knowledge, and beliefs of users), policy and process. Definitions of each construct concept used in this study and how they were mapped onto the final codes are presented in S. Table 2.

## Results

We present the perspectives of 26 respondents from six pilot projects spanning different use cases for DXR-CAD for TB screening and triage. Participants had the following roles: seven project coordinators or managers, three technical or research staff, three radiographers, five radiologists, and eight other clinicians. As these technologies are relatively new, all projects had implemented DXR-CAD for less than a year at the time of interview (Table 1). The themes are presented in terms of facilitators and barriers within the different levels of the CFIR and summarized in Table 3.

**Table 3:** Barriers and facilitators to the implementation of DXR-CAD, presented along the CFIR.

| CFIR Construct(s) | Sub-theme | Facilitator | Barrier |
| --- | --- | --- | --- |
| **Theme 1: Increased portability involves compromises on other device characteristics** |  |  |  |
| Adaptability | Battery and throughput | Various external power sources available | Limited battery life can limit throughout |
|  | Internet dependence | Both online and offline deployment possible | Additional devices required for offline deployment, adding extra weight |
|  | Manoeuvrability & stability | Many options for supporting frames | Heavy-duty frames offer ease of adjustment and better stability, compromising portability |
|  | Image quality & suitability for patients with diverse body sizes | Range of exposure settings to cater for patients of different body size | Some devices cannot be used on large or heavy participants |
| **Theme 2: Complexity increases with integration of hardware and software and interoperable data ecosystem** |  |  |  |
| Complexity | Hardware and software integration | Streamlined integration of products from the same manufacturer/supplier | The integration of combinations of UP DXR and CAD products can be complex when from different suppliers |
|  | Interoperable data ecosystem | Data management system available with some CAD products | Lack of an interoperable and integrated system that would allow the exchange of CAD and DXR data between different health information systems |
| **Theme 3: Limited capacity to implement technologies especially CAD software** |  |  |  |
| Readiness for Implementation | Capacity building | Comprehensive trainings are provided by manufacturers | Online/remote trainings can be less effective |
|  |  | Ongoing support by manufacturers is available after initial training | Manufacturers ill-prepared to offer training on the theory behind the technology, given the opacity of AI |
|  | CAD threshold score selection | Adjustable threshold provides flexibility to adopt different screening strategies | Manufacturer recommended thresholds not always suitable for use-case and setting |
| Knowledge and beliefs about the intervention | Conservative application | WHO recommended that CAD can replace human readers in screening and triaging TB | Insufficient understanding and scepticism on sensitivity of CAD causing delay in behaviour change among clinicians and radiologists |
|  |  |  | Radiologists fear being replaced leading to conservative national policy |
| Theme 4: The lack of guidance on radiation protection suitable for UP XR machines |  |  |  |
| External policy | Radiation Safety Regulation |  | International policy focused on traditional CXR devices that emit more radiation and are used in specialized settings |
|  |  |  | National regulation of radiation safety tends to vary, due to lack of international guidance |
| Theme 5: There is a need to both protect and share patient medical data |  |  |  |
| Executing | Storage and Back-up of Results | Easy to integrate CAD with a range of data storage solutions | CAD hardware can only store a limited amount of data before it inhibits operation |
|  |  | CAD can be configured to automatically back-up to the cloud | Requires at least occasional internet access for back-up |
|  |  | Manufacturers offer de-identifying service prior to cloud upload | Additional costs and hands-on system administration |
|  | Data privacy | A range of de-identification techniques are suitable for CAD interventions | National laws may prohibit transfer of medical data to servers outside the country, limiting options for online CAD deployment |
|  |  | De-identification can occur automatically before upload to commercial clouds | IT expertise may be required to program automatic de-identification |
| Theme 6: Support and maintenance should be responsive to user needs |  |  |  |
| Readiness for Implementation:<br>Available Resources | Maintenance and Support | Comprehensive maintenance packages offered to users | New technology has a propensity to malfunction or for systems to crash |
|  |  | 24/7 online support services | Online support services can only be accessed with internet connection |
|  |  |  | Manufacturers may not have local distributors increasing turn-around time for spare parts |

### Theme 1: Increased portability involves compromises on other device characteristics

Portability was the most frequently mentioned advantage of UP DXR machines, which participants often compared favourably to stationary and heavier-duty mobile X-ray systems.

> *“Regarding the X-ray machine in general, they were quite happy with the portability. One guy took me to the COVID-19 isolation ward and showed me the ‘portable’ X-ray they were using; this was actually a huge X-ray machine and it had to be dragged […] and was […] over 100kg*.*” –* Project Director, Pakistan

Nevertheless, despite marketing claims that the product can be hand-held (the Xair generator) or carried in a backpack (Delft Light), participants indicated that this applied to only some components, while other aspects, such as the detector panel, console and critical accessories, including the supporting frames and lead apron, were not truly “hand-held” or “back-packed” during transportation.

> “*The equipment is not exactly a backpack; the weight of the complete set is nearly 70kg and cannot be operated by only one person in the field… Even these logistics come at an additional cost to us, for example, they might need a motorcycle to take it some places. However, even with this, it is more portable than the truck*.” – Program Coordinator, Nigeria

The miniaturization of the hardware, and its resultant improved portability, came with compromises in meeting the needs of the project from the perspectives of respondents. These compromises included concerns with battery and throughput, internet dependence, manoeuvrability and stability, and image quality and suitability for patients with diverse body sizes; in our analysis, these are categorized as sub-themes and discussed below.

#### Battery and throughput

Limited battery capacity and throughput were mentioned almost unanimously by respondents as the primary disadvantage of using the UP DXR system. The X-ray generator was often mentioned as the rate limiting factor, with the projects in Zambia and Vietnam, who both used the Fujifilm Xair machine, reporting less than 100 shots on one full battery charge. However, other projects received updated versions of the Xair device and reported increased battery capacity. Participants across sites also mentioned that even when the emitter battery was sufficient, another system component, such as the CAD laptop, limited the throughput.

Several users, including all sites using UP DXR for extended periods of time, reported requiring external power sources, such as solar panels and power banks, to increase the number of exposures. However, some participants noted that this adds an extra component and weight, further compromising portability. The acceptability of lower throughput varied depending on the planned intervention aims. One participant noted that while throughput was lower than other equipment, it provided the capacity to offer services in locations they could not reach before, such as primary care level and remote locations:

> “*At first, I was disappointed, because I was coming from using other equipment that could screen 200 patients in a day without worrying about batteries. When we had 20–25 to start with, I was very disappointed. But now I see that with the lower throughput it still has an important utility – as I said, we were able to do screening at the facility level because of it*.” – Project Lead, Zambia

#### Internet dependence

Online or cloud CAD solutions are dependent on strong and stable internet connections, which may not be possible, particularly in remote areas. A slow internet connection can result in a long turn-around-time between image capture and CAD output. As a result, many respondents reported preferring to use offline or hybrid set-ups. A hybrid deployment method allows CAD processing offline, such that results can be synchronized online later, when a fast internet connection is available. Respondents noted that additional hardware is needed in offline or hybrid set-ups, adding weight and complexity to the set-up. For example, at two sites, respondents reported that the laptop with CAD installed provided was heavy, hindering portability.

Using an offline box eliminates the need for a second computer and is more portable, but this option was only available for some UP DXR-CAD combinations, as outlined in Table 1. Furthermore, a router is sometimes needed in addition to the other products, adding yet another component. A fully integrated X-ray system with CAD software – with no need for additional hardware – was not commercially available at the time of pilot implementation or while conducting interviews.

#### Manoeuvrability & stability

Different choices of X-ray generator and detector supporting stands are available and often the more ‘lightweight’ options are offered by default. One respondent radiographer complained of the manual nature of the lightweight generator stand which was tedious when screening people of different heights:

> “*I think the other [problematic] component is the tripod stand. Usually, you discover that you have to alter the stand itself, raise it for the taller person and drop it for the kids. So, there’s this manual aspect of it*.” – Radiographer, Zambia

Another participant switched to heavy-duty tripods, due to the instability of light-weight tripods which makes them prone to falls and damage.

> *“We just switched to more heavy-duty tripods because we had two falls. One caused by wind and the second by an accidental bump*.*” –* Research Director, Vietnam

#### Image quality & suitability of patients with diverse body sizes

Image quality was described as comparable to stationary machines, which can potentially facilitate adoption of UP DXR for case detection. A recent publication from the site in Vietnam showed slightly inferior image quality compared to stationary X-ray, but no effect on CAD interpretation or screening yields [18]. One clinician reported their preference for UP DXR over stationary devices in terms of image quality:

> *“Before we started using [the UP DXR], we had another similar [stationary digital] one […] I think the one that we’re using [now] has better quality images than the one we had before”* – Clinician, Zambia

Just like stationary X-ray, most of the UP DXR systems offered a range of exposure settings for patients of varying size and in different positions. Respondents from one site mentioned taking images of patients lying down who were extremely ill with adequate image quality.

However, this was not the case for all devices. One limitation was related to the Delft Ultra devices, as they cannot be used in patients with a larger body size (above 100kg). As is described by one of the project staff:

“*It produces unclear images when taking the person having a big body or being overweight, although images can still be usable*.” – Radiographer, Ghana

### Theme 2: Complexity increases with integration of hardware and software and interoperable data ecosystem

According to the CFIR, the perceived complexity of implementation is bound up with the duration, scope, radicalness, and disruptiveness of the technologies.^11^ Respondents described levels of complexity in integrating UP DXR and CAD devices, as well as different electronic medical information systems:

#### Assembly and integration X-ray machine and CAD products

The overall assembly process was described as easy after a while, including those that used a heterogeneous product combination. However, one brand of X-ray and CAD, Delft Light with CAD4TB, was noted to be more complicated to set up as it is composed of more devices and wires. The respondents noted to have to develop a packing list to avoid forgetting any hardware before going out to an outreach. Overall, the time required for assembly and disassembly ranged from 5–35 minutes depending on the UP DXR equipment used.

Participants in this study reported using six different combinations of four types of UP DXR and three types of CAD product, as outlined in Table 1. Product combinations from the same manufacturer or manufacturers with a formal partnership (e.g., Delft CXR with CAD4TB and FDR Xair with Lunit INSIGHT respectively) came pre-installed and integrated. However, for participants who used hardware and software from different manufactures, the integration required additional steps whereby both manufacturers were engaged before procurement to ensure that integration was feasible. In the Zambia site, the project lead reported that when using the Fujifilm FDR Xair with CAD4TB another laptop was needed to provide the link between Xair and the CAD4TB cloud. This was because the Xair console laptop was unable to run third-party software (including Google chrome) and so could not access the web browser CAD platform.

Sometimes an additional device was preferred, although not mandatory. For example, when using the Delft Ultra product, although the UP DXR generator included a screen for viewing the CXR result, the poor screen resolution meant the radiographer preferred to use another high-resolution device (either laptop or tablet) to view images.

#### Interoperability of the resulting X-ray screening data with other national health information systems

One of the key challenges was the automatic exchange of CAD and X-ray data between different legacy electronic medical record or health information systems (HIS). Most CAD software providers include an application programming interface (API) that can easily link the X-ray and CAD results to the specific data system of the end-users. However, there is considerable heterogeneity in the data collected, and the HIS landscape is so fragmented that many countries prefer using locally built IT systems. To overcome this challenge, local expertise is needed for mapping of necessary data and database customization (working alongside local IT teams and the CAD and DXR providers).

As a result, diverse solutions emerged. One project site developed a bespoke data collection toolkit and integrated it with the CAD software. Another site used the Excel download from the CAD systems and manually transferred the X-ray CAD data to the national HIS. The other projects only used the locally built screening app or paper-based register and then manually entered the X-ray CAD data.

### Theme 3: Limited capacity to implement technologies especially CAD software

Considering “readiness for implementation”, there was a perceived lack of knowledge and capacity to implement CAD technology among participants, especially regarding setting up and interpreting the threshold score. This despite the manufacturer-led training. In contrast, UP DXR was perceived as easy, partially due to its familiarity as an X-ray device, as opposed to novel CAD technology.

#### Capacity building

Manufacturer-led training and installation of X-ray and CAD was conducted both online (remotely) and in-person for roughly equal proportions of participant projects. Following training, the X-ray process rarely presented difficulty, whereas CAD was seen as challenging, unfamiliar and built on ‘opaque’ concepts and mathematics. Reluctance to bring CAD into clinical decision making was commonly noted. Some users instead relied on clinical evaluation first and then the CAD score:

> “*When I’m interpreting an image, I usually look at the score last. Usually, I keep in my mind what the history and the clinical findings are, then I look at the image itself and what I’m seeing, and if what I’m seeing is highly suggestive of whatever condition I’m thinking of, then I correlate it with the CAD score*.” – Clinician, Zambia

But not everyone is sceptical; some radiographers and radiologists were more open to using the new tool than clinicians, describing it as ‘high-tech’ in a positive way:

> “*I find that using it, the patients make you feel like you’re performing some kind of magic: this is a very simple device, yet making a chest X-ray out of it, you can see the surprise in their faces. It’s just technology doing its work*.” – Radiographer, Ghana

However, some interviewees expressed concerns about healthcare workers using CAD’s prediction alone as the final diagnosis without prescribing any confirmatory testing, as expressed below:

> “*My worry, as it’s rolled out, and as the WHO has now approved it, is that clinicians may interpret a CAD score as TB and I suspect that we already see that […] in some places*” – Program Lead, Zambia

To improve awareness of CAD technology, more theoretical training should be planned, especially for healthcare professionals including not only radiographers but also clinicians, radiologists and decision makers. While there is ample practical training for the radiographers, there is still a lack of knowledge of the theory behind CAD and uses of this technology, which was reflected in some scepticism reported.

#### Conservative application

Regarding the CRFIR construct of beliefs about the intervention, due to lack of capacity and limited understanding of CAD, current application of the technology is tentative in most cases. Caution is reinforced by varying levels of trust in CAD software, due to lack of familiarity, and fears that AI will ‘take over’, particularly among radiologists and clinicians, causing job losses. Only one site described trusting CAD to replace trained radiologists, where it was deployed for rapid triage by the screening camp coordinator. Most implementers were more conservative and required interpretation by a radiologist or other clinicians after CAD:

> “*On CAD you have mixed opinions, certain people really endorse it and get amazed by it, whereas some say you cannot replace a human [with] CAD, you don’t know the knowledge base that has been built up over years of experience*.” – Project Director, Pakistan

Some participants were very impressed with the accuracy of CAD compared to experienced human readers. This, potentially facilitating further scale up of this technology.

> “*Our work shows that there are some software that are really good in our setting and that they perform on par with readers who have over 35 years of experience. I think it’s really nice and I’m hoping we can integrate this as a decision support tool for less experienced radiologists*” – Research Director, Vietnam

The occurrence of perceived false positives and negatives, when compared with radiologist review mostly, was the main reason for questioning the accuracy of CAD decision. This was especially true in those with a history of TB. It is often said that the advantage of CAD is its ability to reduce inter- and intra-reader variability and supplement reader capacity when radiologists are too few, or absent. Our respondents concurred with this but mentioned the need to use their judgement to prescribe treatment.

> “*When someone has had TB before, the score will be high, when we know that the TB has healed and just left a scar, so sometimes we need to use our personal judgement on whether to initiate a person on treatment*.” – Radiographer, Zambia

There were some individual fears of CAD software replacing human radiologists. This may be related to “conservative” national policies which continue to require radiologists be on site where CAD is used:

> *“In general there is hesitancy about the CAD software in Vietnam, the way its communicated in some of the guidelines is that it’s going to replace the radiologists and I think this is at odds with radiologists*.*” –* Research Director, Vietnam

#### Threshold selection

Threshold scores are used to decide if an individual should receive a follow-on diagnostic test or not. Consequently, the score will have a direct impact on sensitivity and the cost associated with follow-on testing. Increasing the threshold score will reduce testing demands (to match available resources and consumables); conversely, reducing the score will mean a more sensitive screening process.

In our study, the issue of threshold score selection strategy tended to divide respondents. A more experienced implementer engaged with the manufacturer to adapt the score in response to low yield. Only at one site did a user describe personally analysing data to select the threshold score (again, one of the most experienced implementers). In contrast, newer implementers seemed more likely to retain the manufacturer-recommended threshold score for some time, until enough confidence, and data, had been gained:

> “*I guess once we finish this study maybe we can edit the threshold and maybe after we’ve done CXR of a good number then we can recommend something*.” – Project Director, Pakistan

One site operated at a threshold score that was selected by the country’s national tuberculosis programme based on TB prevalence screens. Additionally, a clinician reported combining CAD output with their own clinical decision-making:

> *“We don’t strictly stick to 60 and above [threshold], we just use this as a quick judgement when doing population screening. Symptomatic patients with a score of less than 60 are still screened” – Project Coordinator, Nigeria*

### Theme 4: Lack of guidance on radiation protection regulations suitable for UP XR machines

Considering the CFIR construct of “external policy”, there is a lack of international standards on UP DXR. With advances in highly sensitive X-ray detector technology, as well as imaging and noise-reduction technology, certain types of UP DXR can capture high-quality images with low-dose radiation. However, the international guidelines on radiation safety standards for UP DXR have not been updated accordingly: they remain based on standard, heavy-duty X-ray systems, presenting a barrier to implementation in the field.

Due to this lack of international guidance, national guidelines are often followed. In our interviews, respondents describe the national regulations, which vary widely between countries. For example, while one pilot project was allowed to operate outdoor without lead barrier, one project was not allowed to use UP DXR outside at all.

Similarities across country regulations included the use of measurements, being a common requirement for operators to wear dosimeters to measure exposure to radiation. Interviewees indicate that for some of the projects they have conducted measurements of scatter radiation and demonstrated to the local regulatory agencies that the accumulative radiation is much lower than the 1 mSv /year limit set out by the International Atomic Energy Agency [19]. In some instances, investigations were carried out by radiation authorities to approve the devices for use in the field.

In addition to customs clearance, approval from national radiation safety agencies was usually required for the import of X-ray machines into countries; different national radiation authorities have different policies on the approval of importation and use of UP DXR. National radiation safety standards vary; while some national authorities still opted to enforce full radiation safety measures and restrict UP DXR deployment to indoor use, others permitted the use of UP DXR outdoors without a lead wall.

> *“[the National Radiation Authority] didn’t want to give license until the whole room has lead walls, obviously you can’t have this in the field […] They looked at the catalogues, discussed with technicians and everything then they took dosimeters at different distances, 1m, 2m etc when using the machine, and took the dosage at every single place. And at the end they confirmed that if the human body is standing 2m away then they are safe and they can perform without the lead wall*.*” –* Project Director, Pakistan

### Theme 5: There is a need to both protect and share patient medical data

This theme refers to the dual requirements of sharing patient data with other clinicians and at the same time protecting the data to ensure patient confidentiality. There are two sub-themes at play: results storage (including back-up), and data privacy. Both fit under the CFIR concept “process: executing”, which means carrying out or accomplishing the implementation according to plan [17].

#### Results storage and back-up

When using CAD to screen patients, patient data (including the CAD score output and CXR DICOM file) need to be transferred and stored securely. This is essential not only in terms of back-up, but also because CAD laptop storage can swiftly fill up: two respondents reported that this either slowed or prevented the operation of the whole system.

At the projects we surveyed, patient data was either backed up manually (to hard drives, at two sites) or to organizational servers (at one site) or it was automatically uploaded to a commercial cloud for storage (two sites). In one project with low throughput, so such data management systems were required. When using hard back-ups or backing up to the organization server, the implementing organization has more control over the security and privacy of patient data, while uploading to the commercial cloud is an automatic process.

However, the cloud back-up functionality brings benefits such as remote consultation with non-field-based staff to offer a second opinion. The project lead of the project in Zambia described this as one of the main advantages of using CAD.

> *“I felt for us that was very key. When you’re not able to print the X-rays, the radiographers have to be able to log in to the system to see them. Even for support, sometimes I log in from the office to give my opinion on specific X-rays. Sometimes the government radiologists also do this, we just tell them, and they log in and give us an opinion on an X-ray. For us that’s very key*.*” – Project Lead, Zambia*

#### Data privacy

Five sites reported the process of de-identifying data to comply with local data protection or privacy laws. When implementers were using commercial clouds, participants noted that CAD developers were able to facilitate de-identification whereas when implementers were storing data on their own servers, they often already had a system for removing identifying information.

### Theme 6: Support and maintenance should be responsive to user needs

Regarding the CFIR construct “readiness for implementation: available resources”, the respondents reported several hardware faults happened within the first year of usage. Two teams that used Delft Light Backpack and Fujifilm Xair respectively reported the X-ray generator faults six and 11 months after installation. The Delft Light Backpack team also noted a solar panel fault and the long turnaround time to receive the replacement due to lack of in-country stockpile of spare parts. These faults delayed screening activities and service delivery and extra funds were needed to rent generators or other hardware pieces to mitigate the interruption of care delivery.

Additionally, two teams using different UP DXR systems, reported that after a few exposures, the X-ray system heated up, which slowed down its performance. All projects reported positive experiences with the suppliers’ remote support system. However, one of the project staff in Nigeria raised the concern that although suppliers offer 24-hour availability, access to this can be limited for implementers who work in field settings without an internet connection.

## Discussion

This qualitative study identified barriers and facilitators to the implementation of DXR-CAD for TB screening from the perspective of early implementers. The technology was overall perceived as a tool to decentralize TB screening and triage in programmatic settings ranging from facility-based triage to hard-to-reach areas. Early implementers described facilitators such as portability, streamlined integration of products, training and support, as well as the potential to integrate CAD to the health information systems and varied data storage solutions. In contrast, barriers to implementation span the compromises that come with increased portability, namely battery power, internet dependence, manoeuvrability, and image quality for larger patients, the complexity in integrating DXR with CAD systems, limited capacity in implementing CAD, lack of international radiation safety guidance, the need to both protect and share patient data, and the need for comprehensive service and maintenance support.

The context and goals of any intervention will inform the acceptability of the UP DXR. While the relatively limited capacity of the internal built-in batteries with the X-ray system may be a limiting factor, this varies depending on the context of the intended use-case and setting. For example, in primary care facility-based settings, where low daily throughput is expected and electricity supply is expected, these technologies may be ideal. Where very high throughput is important, implementers may consider an alternate power source, supplementary use of a second UP DXR, as was described by one participant, or standard heavy mobile X-ray units, if the setting allows for these additions. In contrast, in very remote areas and where only battery power could be used, population is often dispersed, and small number of participants is often expected. In this case, the ability to access these hard-to-reach populations is the most important factor. Ultimately, balancing the compromises associated to greater portability depends on the goals of the project, and the priorities of the implementer team or program.

One important aspect of integration of CAD is considering the wider health system, and interoperability between national health information systems. The success of the roll-out of new technologies is dependent on an integrated health system alongside the actual technology: new innovations can only be “one part of that solution, necessary but not sufficient” [20]. Thus, to leverage the full capacity of DXR-CAD, enabling this interoperability between data systems is key. The digital nature of these tools presents an opportunity for the harmonization of digital ecosystems for TB surveillance. Most CAD software providers include an application programming interface (API) that can easily link the X-ray and CAD results to the specific data system of the end-users.

While most countries have a surveillance system, the high heterogeneity in the data collected, and the considerably fragmented HIS landscape pose challenges to follow the entire screening cascade and ensure TB treatment initiation (such as DHIS2 or eTB Manager) [21]. Measures to overcome this can include, but are not limited to, setting up basic standardized TB health data formats and elements and developing necessary API to allow data flow between systems. Local expertise is also needed for data mapping and database customization. CAD developers should expect to work with countries to develop workable and scalable models for key information exchange and adaptation to different countries.

Advances in UP DXR technology were accepted, perhaps because they build on radiological knowledge already well-established. In contrast, respondents revealed a mixed picture of the general acceptability of CAD technology, particularly due to fears of replacement and a poor understanding of software’s underlying algorithms. These findings align with a recent survey showing that while healthcare workers in the UK generally think artificial intelligence could be useful in medicine, they lack a full understanding of its underlying principles and are worried about potential consequences of its widespread use [22]. Current CAD training practices generally fail to address important individual, cultural or organizational assumptions that underlie attitudes to the technology.

These barriers are similar to described in experiences with other novel technologies; for example, scepticism among healthcare workers was reported when rolling out Urine LAM tests for diagnosis despite promising evidence [23]. The heterogeneity of feedback about CAD in this study could relate in part to the CFIR concept “tension for change” – the degree to which stakeholders perceive the current situation as needing change [17] – in this case the availability of radiologist readers. It is worth noting that clinicians and radiologists may be critical of CAD’s detection of false positives and negative, without critically considering that human readers are prone to similar errors. A clinician’s belief in their own capabilities to use CAD to make referral decisions (self-efficacy) can also determine how open they are to behaviour change using CAD. Furthermore, considering fears that CAD may replace radiologists, more could be done to evaluate use cases where CAD software is used as a decision support tool alongside human readers, rather as a replacement.

Finding an optimal threshold score for a given setting is presented a barrier to implementation for several respondents. Manufacturers sometimes propose a default threshold; however, as outlined by the WHO, manufacturer-recommended threshold scores are not always the most appropriate for the setting and use case and can result in unexpected and undesired false negatives and false positives [24]. Furthermore, some manufacturers do not propose a threshold, leaving implementers to choose. In these cases, and if it is not feasible or practical to conduct proper operational research to select the most scientific threshold score associated with a designated sensitivity or specificity before implementation, then the implementation should be accompanied with monitoring yield, screen positive rate, positive confirmation test rate among other factors, to then adjust the threshold accordingly. A standard operating procedure is needed to articulate and define the steps to change the threshold score. For example, an iterative operating point calibration (ITSC) mode was signed that allow TB programs to start with a rough threshold score and refine it through iterative cycles with new data during implementation until programmatic targets are reached [25]. By providing the opportunity to refine the threshold score in parallel to an ongoing intervention, ITSC is suitable for sites where time and resources do not allow for suitably rigorous research at the outset of an intervention.

### Strengths and limitations of the study

This study is the first to assess the early users’ experience of DXR-CAD technologies for TB. This is partly a limitation because these technologies are new, and the respondents can only describe their own relatively brief experience during a pilot phase. Perceptions and acceptability may change as implementers become more familiar with these tools. Similarly, due to rapid change, some of the issues highlighted by respondents have now been improved by manufactures. Respondents from one project (Ghana) are affiliated with the manufacturer of the device, which is very new on the market. However, this was not the case for the remaining respondents.

### Outlook for future implementers

There are several decisions that policymakers, country programmes and manufactures can take to ensure the successful implementation of these technologies. Firstly, there is a need to update international radiation safety standards to keep abreast of technological advances behind new X-ray devices with much lower radiation and higher dosage efficiency. Current international radiation safety standards should not constrain this technology. Given that any significant changes to international policy takes time, it is encouraging to see that some national regulatory agencies are taking bold initiatives to introduce this technology, with stringent monitoring plans in place to ensure safety; this alongside more data on radiation exposure as the use of the technology increases may lead to changing attitudes towards the safety of these devices.

The implementation of this technology is evolving rapidly, but several areas are clearly still open to improvement, especially the integration of X-ray machines and CAD software. In the projects surveyed, no CAD software can be installed directly on the X-ray console laptops. As a result, additional devices and connecting leads are required, particularly when the hardware and software are provided by different suppliers and for offline deployment. Software developers should partner with X-ray equipment manufacturers to further streamline the integration to reduce the complexity of assembly and the consequent transportation burden for users. Manufacturers of certain ultra-portable DXR systems should reduce the number of wires and hardware pieces to reduce the overall complexity of set up or assembly to improve user-friendliness.

Countries and programs thinking of using CAD technology should plan and budget for backing up and synchronizing the data ahead of time. As pilot projects are scaled up to national rollout, data, back-up and synchronization will become a bigger issue alongside issues relating to data privacy and security. Data privacy and security are clearly essential to protect the human rights of people affected by TB and others who provide their personal health data for processing using CAD technologies. Although this is of fundamental importance, it need not to present a problematic hurdle. Safeguarding data privacy and security can and should be achieved without interfering with the use and functioning of CAD technologies. Implementers can use a range of legal, technical, and operational measures to safeguard the collection, storage, and processing of data while still ensuring the full functionality and benefit of CAD technologies.

Further, it is recommended that users should purchase a comprehensive and long-term warranty and support package [26]. The technologies are new and there have been malfunctions reported by these pilot projects interrupting screening activities and sometimes incurring extra costs, as outlined in our results. As the technologies are so new, early implementers have a role to play in advocating for product and service improvements by manufacturers, based on their on-the-ground experiences. We hope that our manuscript presented here will also encourage manufacturers to tailor their products to meet the needs of end-users and that the hardware and software will become more robust as the manufacturing matures with time.

### Conclusion

This qualitative study has found that the respondents were able rapidly to use UP DXR and CAD following initial training, while the increased portability provide a great opportunity to decentralize radiological assessment. Further streamlining of the integration of X-ray and CAD equipment from different manufacturers and with different data would greatly enhance the user experience and reduce complexity. Two of the main barriers to uptake are the lack of understanding of CAD software – particularly threshold score selection – and the lack of international and national guidance on radiation protection suitable for UP DXR. Programmatic planners should seek guidance from their national radiation authority as early as possible and incorporate training to ensure capacity. Safeguarding data privacy and security can and should be achieved without interfering with the use and functioning of CAD technologies. Finally, programmatic planners should include comprehensive warranty and support packages.

## Data Availability

Data cannot be shared publicly. The personal information of the participants and all other confidential information has been stored and secured according to the medical confidentiality and other provisions of the General Data Protection Regulation (GDPR), the German Federal Data Protection Act (BDSG) and the associated Baden-Württemberg State Data Protection Act (LDSG BW).

## Acknowledgements

The authors would like to acknowledge the contribution of all the respondents: Drs Monde Muyoyeta, Charles Imbuwa, Brian Shuma, S N, Vo Nguyen Quang Luan, Andrew Codlin, Kinz-Ul-Eman, Maurits Verhagen, Nicholas Setor Korto, Austin Ihesie, Bethrand Odume, Eze Chukwu, Kim Hee Jin, Kim Ji Yun, Kim Su Hee.

## Authors’ contributions

ZZQ and RB conducted pilot interviews. ZZQ, RB, and MMC developed and applied the coding scheme supervised by CDM and JC. ZZQ led the interview. RB, MMC, SZ assisted in coding analysis, applying CFIR framework and formulate the themes with significant contribution and supervision from CDM and JC. ZZ drafted the first manuscript with the assistance from RB and SZ. MMC, AJC, CDM and JC provided critical review and helped with the revision of the manuscript. All authors read and approved the final manuscript.

